# Long COVID Disability Burden in US Adults: YLDs and NIH Funding Relative to Other Conditions

**DOI:** 10.1101/2024.01.09.24301057

**Authors:** Karen Bonuck, Qi Gao, Seth Congdon, Ryung Kim

## Abstract

**Background:** Long COVID (LC) is novel, debilitating and likely chronic. Yet, scant data exist about its disability burden to guide scientific research and public health planning. We estimated Long COVID’s non-fatal disease burden in US adults and its FY2024 actual: burden-commensurate research funding from the National Institutes of Health (NIH) relative to other conditions, and biological sex.

**Methods:** We present YLDs/100,000 for 70 NIH Research, Condition, and Disease Categories (RCDCs). Prevalence of disabling Long COVID was obtained from cross sectional surveys of representative samples of US adults, from September 2022 to August 2023. Disabling Long COVID was defined as incident symptoms persisting more than 3 months post-COVID, that significantly compromise daily activities. We calculated burden-commensurate funding for the top YLD conditions and for female vs. male dominant conditions.

**Findings:** Disabling Long COVID was reported by 1.5% (n= 10,401) of n=757,580 respondents: Compared to the overall sample, those with disabling LC disproportionately identify as female (64.4% vs. 51.4%) and experiencing disability (80.8% vs. 52.9%) anxiety (57.5% vs. 23.8%) and depression (51.3% vs.18.5%). It ranked in the top 25% of YLDs at 320/100,000, between Alzheimer’s (279.4/100,000) and asthma (355.7/100,000) but received just 10% of its actual: YLD-commensurate funding. Only 5 conditions received *less* actual: burden: commensurate funding, including Myalgic Encephalitis/Chronic Fatigue Syndrome (<1%), another post-viral, female-dominant condition.

**Interpretation:** LC has debilitated 3.8 million (weighted frequency) US adults. Research funding for it, like other female dominant conditions, lags behind its disability burden.

**Research in Context:** <u>Evidence before this study</u> – We analyzed Long-COVID’s (LC) non-fatal disease burden in the US--represented by YLD (years lived with disability= prevalence x disability weight) -- and National Institutes of Health (NIH) research 2024 funding relative to other conditions. We searched PubMed through 11/28/2023 for Long COVID prevalence (US), and Long COVID disability and disease burden (not US-specific). The keywords “years lived with disability” + “COVID” yielded n= 38 articles (11/29/23); but most referenced “disability-adjusted life years” (DALYs) in other countries. Similarly, “disease burden” + Long COVID yielded 23 papers, but no US YLD data. See Supplement 1 for meta-analyses, systematic reviews and US studies of Long COVID prevalence and impact.

We instead sourced YLD data from the US Census Bureau’s Household Pulse Survey (HPS) and the Institute for Health Metrics and Evaluation (IHME) /Global Burden of Disease (GBD) Long COVID Study Group. The HPS queries adults about Long COVID-related symptoms and their impact on daily activities. We applied the IHME/GBD’s estimated Long COVID disability weight of 0.21 and harmonized it with our LC case definition from the HPS data in consultation with IHME/GBD researchers. To harmonize IHME/GBD disability weights for *non-LC* diseases/conditions with the NIH’s terminology, we consulted with NIH staff. LC definition and measurement affects prevalence and burden estimates; our use of high-quality data sources and transparency in reporting how they were applied reduces the risk of biased assumptions.

<u>Added value of this study-</u> Long COVID is a chronic debilitating condition. While there is ample research on COVID’s acute illness and loss of life, there are no population-based data on its disability burden. We provide that data. To guide scientific research and public health planning, we report YLDs associated with disabling Long COVID (i.e., symptoms significantly limit activity), and; compare it to other conditions’ YLDs, NIH funding, and female-vs. male-dominance. It ranked in the top 25% of YLDs at 320/100,000, between Alzheimer’s (279.4/100,000) and asthma (355.7/100,000) but received just 10% of its YLD-commensurate funding. Only 5 conditions received less burden-commensurate funding; 3/5 were female-dominant, including Myalgic Encephalitis/Chronic Fatigue Syndrome (ME/CFS) at <1%, another post-viral condition that shares significant overlap with Long COVID. Overall, median funding/YLD was >= 5 times greater for male-vs. female-dominant conditions.

<u>Implications of all the available evidence</u>-Nearly 4 million US adults (weighted frequency) live with disabling Long COVID. They disproportionately identify as female and as having a disability, anxiety and depression. Yet NIH funding for diagnostic and treatment research for Long COVID hasn’t kept pace with its disability burden.

## Introduction

Long COVID (LC) is a multi-systemic condition that affects virtually every organ with its 200+ associated conditions^1–3^ that commonly cluster into fatigue, respiratory and/or cognitive phenotypes.^4^ Long COVID occurs in 10%-30% of people after acute infection^3–10^ depending on age^5,11^and severity of infection,^5^ and is known to persist for at least one^7^ to two^9^ years post-infection. Women, people of color, those who are LGBTQ and/or had pre-existing disabilities are also disproportionately impacted.^1,3,6,12^ Long COVID currently lacks validated diagnostic markers and treatment protocols.^1,3,13,14^ Further, standard testing routinely yields unremarkable findings despite clear physiological dysfunction.^1,15^ Consequently, patient concerns are often dismissed as psychosomatic-which leads to diagnostic and treatment delays.^1,14,15^

Long COVID represents a mass disabling event of significant public health concern.^1,16,17^ Long COVID is associated with a 21% loss of health – comparable to traumatic brain injury or complete hearing loss.^18^ Among US adults, 5.3% reported having Long COVID in October 2023 and 1-in-4 with Long COVID consistently report *significant* activity limitations from its symptoms.^19^ Without effective treatments on the immediate horizon, advocates and scientists have criticized the pace, scope and focus of US-funded clinical trials.^15,20^ In fact, just n= 33 Long COVID (or “PASC”) intervention studies were listed in <u>ClinicalTrials.gov</u> as of 5 December 2023 [seeking + no longer seeking participants; both sexes; all ages; no date limit]. Only one was NIH-funded, in contrast to NIH funding 399 of 1,039 HIV/AIDS intervention trials (date limit: 1/1/2020-12/5/2023). NB-Terms for long COVID include: PASC (Post-Acute Symptoms of COVID-19), PCC (Post-COVID Condition) and long-haul COVID. This paper uses the term Long COVID.

Burden of disease (e.g., prevalence, mortality, morbidity, disability) is assumedly a significant factor in NIH funding decisions, along with health care costs, scientific opportunity, and disease-specific advocacy. However, analysis of NIH funding from 2008 to 2019 found that neither burden of disease--nor changes in burden of disease--was a significant factor. Instead, the strongest predictor was prior funding, which led researchers to surmise “…that changing public health needs have a limited role in guiding NIH funding.”^21^ Likewise, leading Long COVID researchers observed no change in apportionment of NIH funds *outside* the pandemic vs. *in* the midst of it.^17^ Long COVID, with its multiple causes and diverse phenotypes,^4,17^ precludes a simple single cure. Seemingly, such a condition affecting millions in the US, should drive substantial NIH investment.

Long COVID’s clinical characteristics overlap with Myalgic Encephalomyelitis/Chronic Fatigue Syndrome (ME/CFS) and dysautonomia (particularly its postural orthostatic tachycardia syndrome [POTS] sub-type);^1,2,13,16,22^ risk of these conditions increases by up to 60% and 80%, respectively, after COVID infection.^1^ The three conditions: have shared etiology along with multidimensional, episodic and disabling symptoms; lack treatment protocols and; have uncertain prognoses. ^1–3,13,15^ Further, all are female-dominant conditions, generate psychological distress, and; are often dismissed as psychosomatic.^1–3,13,23^

The NIH funds ME/CFS at just 7.3% of its commensurate burden measured in DALYs (disability-adjusted life years);^24^ there is no analysis of dysautonomias’ burden-commensurate research funding. In nearly 75% of cases where a disease is female-or male-dominant, NIH funding favors males, i.e., disease affects more women but is underfunded (relative to burden) or disease affects more men and is overfunded (relative to burden).^24^

Long COVID is a novel, debilitating and likely chronic condition for many— based on lived experience^15^ and how chronic conditions are generally defined: >=12 months duration, and; requires ongoing care and/or limits daily activities. Yet, despite millions of US adults living with Long COVID, the empirical population-based data needed to guide scientific research and public health planning, are lacking. This study fills that gap by: quantifying the non-fatal disease burden of <u>disabling</u> Long COVID in the US; comparing it to other conditions and actual: burden-commensurate NIH funding for them, and; comparing female-vs. male-dominant conditions by their actual: burden-commensurate funding.

## Methods

### Description of Measures

*NIH Funding* – We use data from the NIH’s *Estimates of Funding for Various Research, Condition and Disease Categories* (RCDC) report which provides prevalence, mortality, and DALYs for ≈75 RCDCs (link: https://report.nih.gov/funding/categorical-spending<u>#/</u>.) We included these non-LC comparator conditions (except for ME/CFS, see below) from the RCDC because there are IHME/GBD Study-derived disease burden measures for them.

*Long COVID Disease Burden* – We use YLD (“years lived with disability”) to measure the non-fatal disease burden of debilitating Long COVID. We chose YLD over other burden metrics, e.g., DALYs or YLL (years of life lost to disease) because a) disability— not death— now accounts for most COVID burden,^25^ b) Long COVID has evolved into a chronic condition for many, for which YLD is the optimal measure,^5,7–9^ and; c) lack of longitudinal data needed to estimate YLLs, especially at younger ages. YLD is calculated as: *YLD= prevalence x disability weight*

*Long COVID Disability Weight*-Disability weights represent health loss from non-fatal diseases, where 0= full health and 1=death. The IHME/GBD study produces mortality and disease burden (i.e., YLD, YLL, DALY) data from epidemiologic datasets, population surveys, medical records and the scientific literature. (See: https://www.healthdata.org/research-analysis/gbd). They published disability weights for three common symptom clusters following symptomatic COVID-19 in 2020 and 2021: persistent fatigue, cognitive, and respiratory,^4^ and estimate Long COVID’s disability weight as 0.21.^13,26^ This weight--confirmed by IHME/GBD researchers (Sarah Wulf Hanson email to author, 8/3/2023) (Theo Vos emails to author, 8/3/3023 and 1/8/2024)—is slightly lower than the 0.23 from the latter reference^26^ as new studies were added to the estimate (Theo Vos email to author, 1/8/2024).

*Non-Long COVID Disease Burden*-We applied the IHME/GBD’s YLDs for adults >=20 years to the RCDCs that NIH had harmonized with IHME/GBD categories (link: https://report.nih.gov/report-nih-funding-vs-global-burden-disease). In the rare cases when >=2 IHME/GBD categories were combined to match an RCDC category, their disease burdens were summed. [Bochner D, National Institutes of Health, emails on 7/17/2023 and 12/1/2023]. See Supplement Table 3 for RCDC categories, their YLDs, and NIH 2024 funding. Of the RCDC conditions -- including LC and ME/CFS (see note below), 6 were excluded from analysis: 5 because harmonizing the IHME/GBD and RCDC categories was difficult and thus we couldn’t extract their YLDs (mental illness, heart disease, Injury (total) Accidents/Adverse Effects, cancer, and sudden infant death syndrome), and 1 because its YLD= 0 (Malaria).

Note-We included ME/CFS into our analysis, despite it not being in the IHME/GBD dataset, given its clinical overlap with disabling Long COVID. To estimate the YLD of ME/CFS, we used recent US prevalence data [total= 1.3%, women= 1.7%, men= 0.9%]^27^ and a disability weight [0.46] from prior research.^23,28^)

*Non-Long COVID Disability Weights* (**Figure 1**)-For context, we present the disability weights of Long COVID and selected chronic conditions, from the IHME/GBD Study [see: www.thelancet.com/cms/10.1016/S0140-6736(20)30925-9/attachment/7709ecbd-5dbc-4da6-93b2-3fd0bedc16cc/mmc1.pdf Table S13. (p. 1528)]. The disability weight of Long COVD is roughly equivalent to that of traumatic brain injury (0.21), complete hearing loss (0.22) and moderate chronic obstructive pulmonary disease (0.23). It exceeds that of uncomplicated diabetes (0.05) and mild Alzheimer’s (0.07), and is exceeded by symptomatic HIV (0.27), moderate rheumatoid arthritis (0.34) and migraine (0.44).

**Figure 1:**
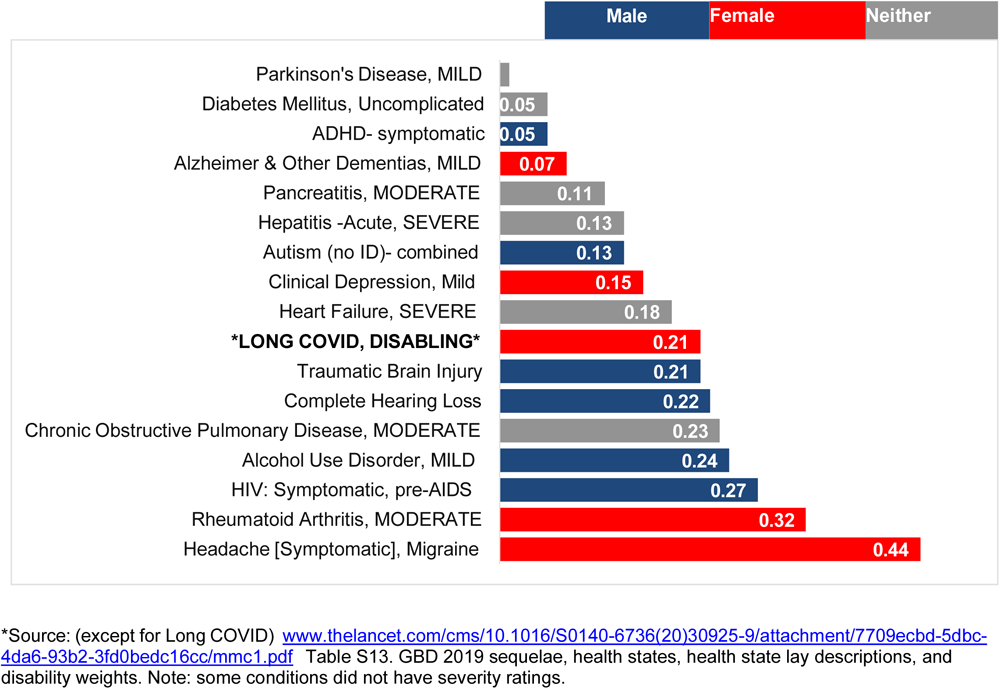
Disability Weights of Disabling Long COVID vs. Selected Conditions, by Female-, Male-or Neither-Dominant Condition.

*Long COVID Case Data Source*-The Census Bureau’s monthly online Household Pulse Survey (HPS https://www.cdc.gov/nchs/COVID19/pulse/long-COVID.htm) from 9/2022-8/2023 was used to identify cases. This monthly repeated cross-sectional study measures COVID’s impact on US households from a representative sample of adults (>= 18 years). To harmonize the HPS and IHME/GBD data, we excluded HPS participants <20 years.

*Long COVID Case Definition*-The HPS and IHME/GBD both operationalize Long COVID as incident symptoms >= 3 months post-infection, but they differ in the symptoms they include (see Supplement Table 2). Per above, the LC disability weight of 0.21 is derived from 2020-2021 data for symptoms following symptomatic disease (i.e., excluding asymptomatic COVID). The HPS symptoms, while less severe than those in the IHME/GBD study, match the symptoms listed in the July 2021 federal guidance designating LC as a qualifying disability in the US https://www.hhs.gov/civil-rights/for-providers/civil-rights-covid19/guidance-long-covid-%20disability/index.html#:~:text=Can%20long%20COVID%20be%20a,or%20more%20major%20life%20activities

To harmonize differences between the HPS and IHME/GBD symptom descriptions, we restricted our analysis of Long COVID YLDs to just HPS respondents reporting significant activity limitations from Long COVID symptoms. For this case definition respondents had to answer “Yes” to the first three questions below and “Yes, A Lot” to the fourth question:

1. Have you ever tested positive for COVID-19 (using a rapid point-of-care test, self-test, or laboratory test) or been told by a doctor or other health care provider that you have or had COVID-19?
2. Did you have any symptoms lasting 3 months or longer that you did not have prior to having coronavirus or COVID-19? See Supplement Table 2.
3. Do you have symptoms now?
4. Do these long-term symptoms reduce your ability to carry out day-to-day activities compared with the time before you had COVID-19? (Options: “No, not at all,” Yes, a little” and “Yes, a lot.”)

*Long COVID Prevalence*-- For each month, the point prevalence estimate (per 100,000) of Long COVID with limiting symptoms were calculated using 80 replicate survey weights provided by the HPS. The standard errors were calculated by jackknife method. Then the point prevalence estimates of 12 months were averaged as the annual point prevalence estimate.

*Sex-Dominant Diseases/Conditions*- If a disease or condition affected at least 60% of one biological sex, we classified it as female or male dominant, per prior work.^24^ The remainder were identified as neutral.

*Demographics*- We describe the aggregate 12-month HPS sample by Long COVID status as a) Current LC-Activity Limit: No/Little, b) Current LC Activity Limit: A Lot (i.e., disabling) c) Past LC, and; d) Other-never COVID and never LC. Disability status was based on having (any) difficulties seeing, hearing, walking or climbing stairs, remembering or concentrating, doing daily activities and/ or communicating. To ascertain mental health status, the HPS incorporated the 2-item Patient Health Questionnaire (for depressive symptoms) and the 2-item Generalized Anxiety Disorder scale.

### Statistical Methods

The frequency and annual point prevalence (number of cases per 100,000) of Long COVID among the adults (age>=20) were estimated using the survey weights. Specifically, 12 monthly survey data were combined with their replicate and base survey weights divided by 12. The YLD of Long COVID was calculated by multiplying the prevalence with the disability weight of 0.21

A scatter plot was generated to visualize the possible association between the NIH 2024 estimated funding and the YLDs with logarithmic scale on both axes. Basic descriptive statistics for patient characteristics were reported as frequency, weighted frequency and percentage calculated from the pooled data set by using the method for calculating the prevalence of Long COVID. The commensurate funding for each condition was estimated by multiplying the total funding of the 70 conditions with the proportion of its YLD among the total YLDs. The funding per YLD was calculated by dividing the funding for each condition by its YLD. The Wilcoxon rank sum test was used to compare the difference of the funding per YLD between male and female dominant conditions.

## Results

### Sample Characteristics by LC Status *(Table 1)*

Across the 12-month sample of n=757,580 US adults, 1.5% (n= 10,401) met our case definition of disabling Long COVID. Their estimated frequency of the population equates to 3,801,986 adults with long term symptoms after COVID that significantly limits daily activity. Compared to the overall sample, they disproportionately identify as female (64.4% vs. 51.4%) and experiencing a disability (80.8% vs. 52.9%), anxiety (57.5% vs.23.8%), and depression (51.3% vs.18.5%). An additional 4.5% (n= 35,963) had Long COVID with little or no activity limits. Thus, altogether > 15 million US adults report Long COVID. The prevalence of debilitating Long COVID stabilized after December 2022 (not shown), as reported by others.^19^ The proportions receiving a(ny) COVID vaccine or booster did not appear to differ by Long COVID status.

**Table 1:** Sample Characteristics by Long COVID Status.

|  |  | TOTAL | Current LC:<br>Activity Limit=<br>A LOT (Disabling) | Current LC:<br>Activity Limit=<br>NO/LITTLE | Past LC:<br>Not currently | ELSE |
| --- | --- | --- | --- | --- | --- | --- |
| # Respondents |  | 757,580<br>(100%) | 10401<br>(1.5%) | 35963<br>(4.5%) | 59294<br>(8.5%) | 651922<br>(85.5%) |
| Estimated Frequency,<br>All US Adults |  | 249,590,271 | 3,801,906 | 11,231,229 | 21,278,202 | 213,278,933 |
| **Age:** | 20-29 | 64102<br>(15.6%) | 643<br>(12.2%) | 2816<br>(15%) | 6570<br>(19.3%) | 54073<br>(15.3%) |
|  | 30-39 | 139104<br>(18.8%) | 1638<br>(16.1%) | 6776<br>(19.3%) | 13384<br>(22.4%) | 117306<br>(18.4%) |
|  | 40-49 | 143581<br>(17.2%) | 2373<br>(21.3%) | 8234<br>(20.4%) | 13624<br>(20.5%) | 119350<br>(16.6%) |
|  | 50-59 | 135907<br>(17.1%) | 2499<br>(21.3%) | 7745<br>(19.9%) | 11626<br>(17.8%) | 114037<br>(16.7%) |
|  | 60-69 | 154479<br>(18.6%) | 2156<br>(18.9%) | 6749<br>(17%) | 9297<br>(13.7%) | 136277<br>(19.1%) |
|  | 70+ | 99182<br>(10.4%) | 1092<br>(10.2%) | 3643<br>(8.4%) | 4045<br>(5.3%) | 91176<br>(11.2%) |
| **Sex assigned at birth:** | Female | 427865<br>(51.4%) | 7267<br>(64.4%) | 24976<br>(64.8%) | 39910<br>(61.6%) | 355712<br>(49.5%) |
| **Education:** | <High School Diploma | 15995<br>(7.4%) | 390<br>(12.4%) | 617<br>(5.2%) | 1512<br>(8.1%) | 13476<br>(7.3%) |
|  | HS Diploma or GED | 92913<br>(30.2%) | 1480<br>(29.4%) | 4022<br>(26.3%) | 8202<br>(30.5%) | 79209<br>(30.4%) |
|  | Some college/Associate | 239562<br>(29.7%) | 4611<br>(37.2%) | 13822<br>(36.8%) | 22238<br>(33.7%) | 198891<br>(28.8%) |
|  | BA or higher | 409110<br>(32.7%) | 3920<br>(21.1%) | 17502<br>(31.7%) | 27342<br>(27.6%) | 360346<br>(33.5%) |
| **Disability status:** | Yes (vs. No) | 414289<br>(52.9%) | 8789<br>(80.8%) | 27221<br>(74.1%) | 38274<br>(61.7%) | 340005<br>(50.4%) |
| **Ethnicity:** | Non-Hispanic<br>(vs. Hispanic) | 691427<br>(82.7%) | 9198<br>(78.2%) | 32599<br>(83.1%) | 51972<br>(76.6%) | 597658<br>(83.4%) |
| **Race** | White | 621230<br>(75.5%) | 8334<br>(75.5%) | 30247<br>(79.6%) | 48295<br>(76.3%) | 534354<br>(75.2%) |
|  | Black | 61250<br>(12.5%) | 791<br>(10.3%) | 2445<br>(9.5%) | 4850<br>(11.5%) | 53164<br>(12.8%) |
|  | Asian | 36109<br>(5.7%) | 222<br>(3%) | 916<br>(3.4%) | 2141<br>(4.5%) | 32830<br>(6%) |
|  | Others | 38991<br>(6.3%) | 1054<br>(11.1%) | 2355<br>(7.4%) | 4008<br>(7.7%) | 31574<br>(6%) |
| **COVID Vaccine:** | Ever<br>(vs. Never) | 653675<br>(82.7%) | 8785<br>(80.7%) | 31848<br>(86.1%) | 50454<br>(82.6%) | 562588<br>(82.6%) |
| **Anxiety:** | Yes<br>(vs. No) | 167280<br>(23.8%) | 6023<br>(57.5%) | 13043<br>(38.5%) | 18979<br>(33.1%) | 129235<br>(21.6%) |
| **Depression:** | Yes<br>(vs. No) | 122140<br>(18.5%) | 5217<br>(51.3%) | 9598<br>(29.5%) | 13731<br>(25%) | 93594<br>(16.7%) |

### Top YLDs/100,000 by Female-, Male-or Neither-Dominant Condition *(Figure 2)*

This figure presents the n=18 conditions in the top 25% of YLDs/100,000, i.e., the highest magnitude of non-fatal disease burden in US adults. Long COVID was 16^th^ out of the 70 conditions at 320/100,000, just above Alzheimer’s (279.4/100,000) and Alcoholism, Alcohol Use and Health (303.7) and below asthma (355.7) and schizophrenia (397.5).

**Figure 2:**
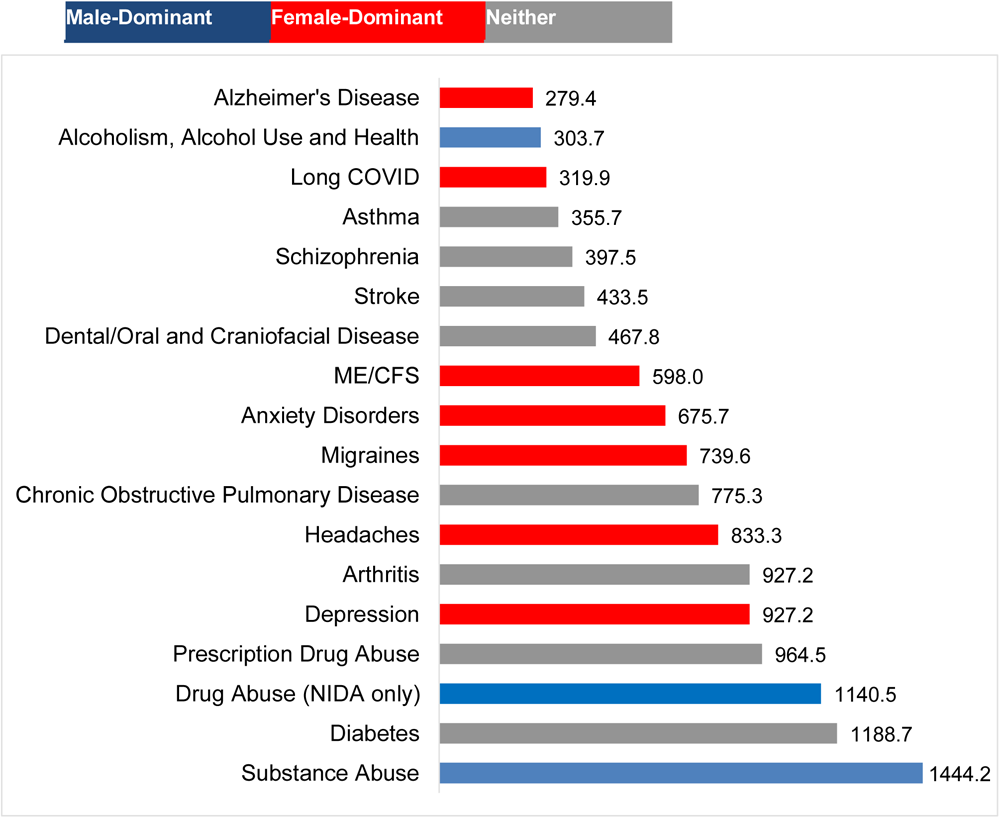
Top n=18 YLDs/100,000 by Female-, Male-or Neither-Dominant Condition.

### YLD vs. NIH Funding by Female-Male-or Neither-Dominant Condition *(Figure 3)*

We visualized the 70 conditions by YLDs/100,000 (X axis), and NIH 2024 funding in millions of US$ (Y axis). See Supplement Table 3 for exact YLD and funding data. The scatterplot is bisected at the vertical median for YLD at 48.4 and at the horizontal median at $275.5 million. Long COVID appears in the lower-right quadrant with 13 other conditions that are under-funded relative to YLDs. Of the quadrant’s 14 conditions, 8/14 are female-dominant, none are male-dominant (Anxiety received $274 million, just below the median).

**Figure 3:**
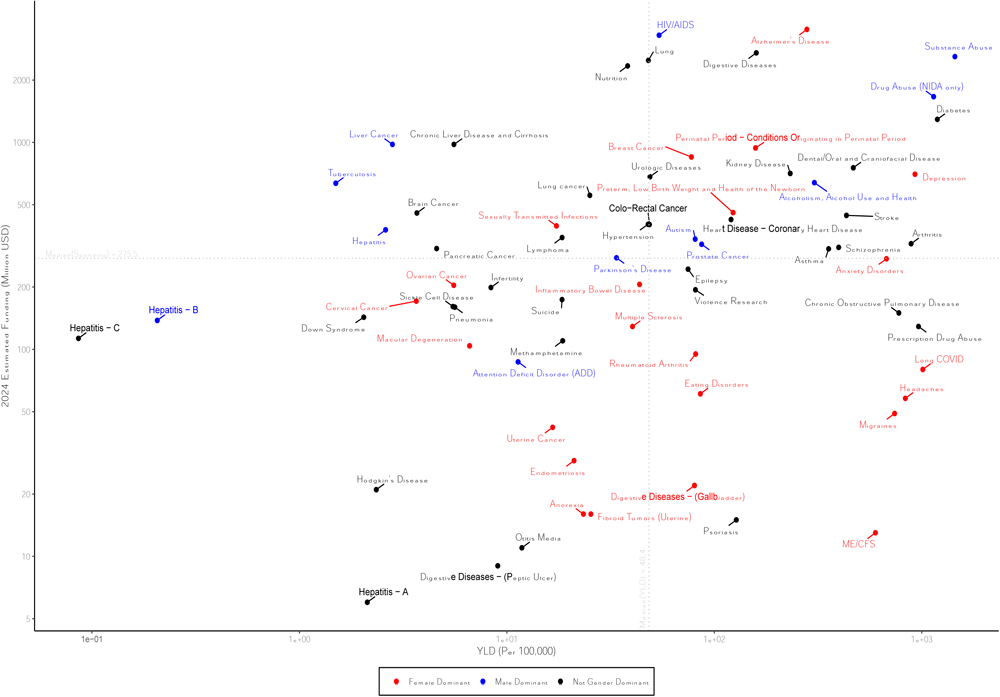
YLDs/100,000 by NIH 2024 Funding (Millions, USD) for 70 Conditions in US Adults.

### Burden-Commensurate Funding for Top n=18 YLD Conditions by Female-, Male-or Neither Dominant Condition *(Figure 4)*

Long COVID research was funded at 10% of its actual: burden-commensurate amount. Among the top 25% conditions by YLD (n= 18), just five have lower actual: burden-commensurate funding -- ME/CFS (1%), Migraines (3%) and Headaches (3%)—all female-dominant like Long COVID. In contrast, the most over-funded condition was female-dominant Alzheimer’s, at 498%, followed by two male-dominant conditions, Alcoholism (83%) and Substance Abuse (71%),

**Figure 4:**
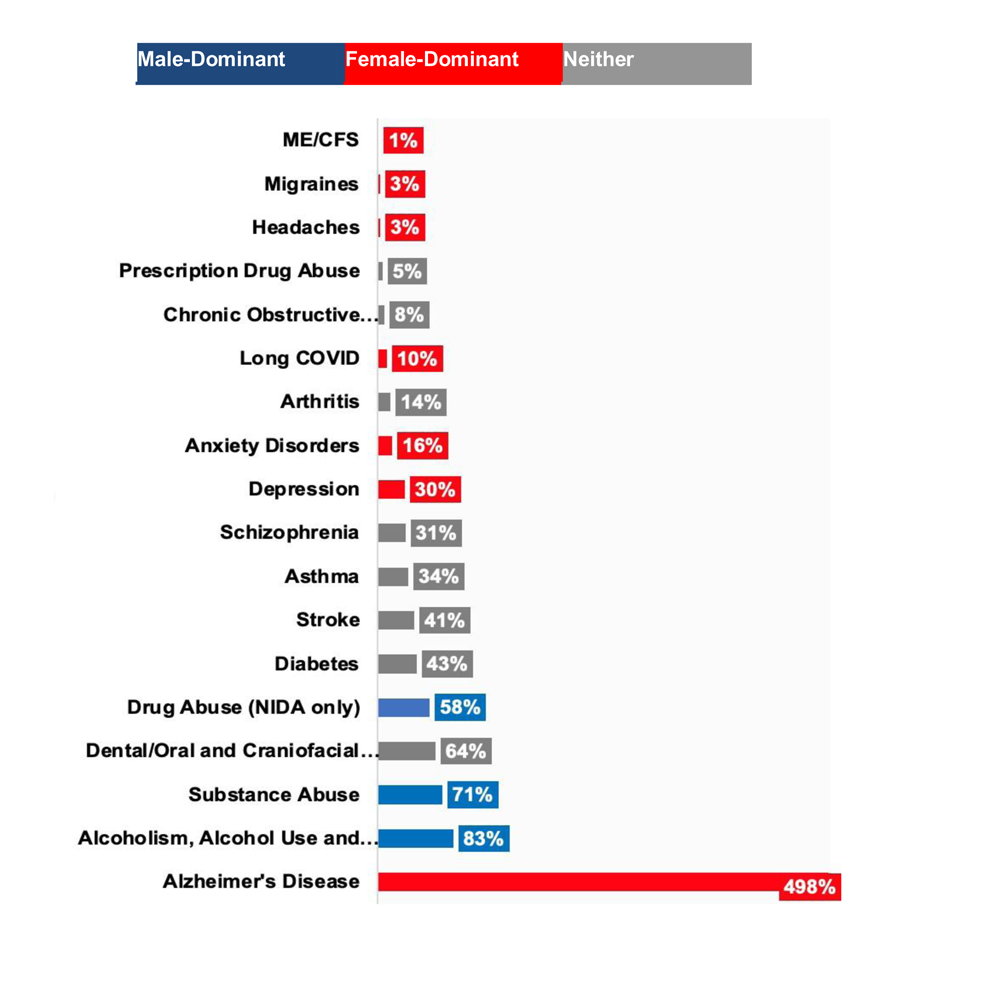
Actual: Burden-Commensurate Funding for Top 18 YLD Conditions by Female-, Male-or Neither Dominant.

### Funding per YLD: Male, Female or Neither Sex-Dominant Conditions *(Figure 5)*

Conditions classified as male-(n= 12) vs. female-(n= 23) dominant received 5-fold more median funding per YLD, (7.9 [IQR: 1.0-22.6] vs. 1.4 [IQR: 0.5-8.5], p=.009). Comparatively, median funding per YLD was higher for conditions not dominant by either sex (8.2 [IQR: 1.1-26.1])

**Figure 5:**
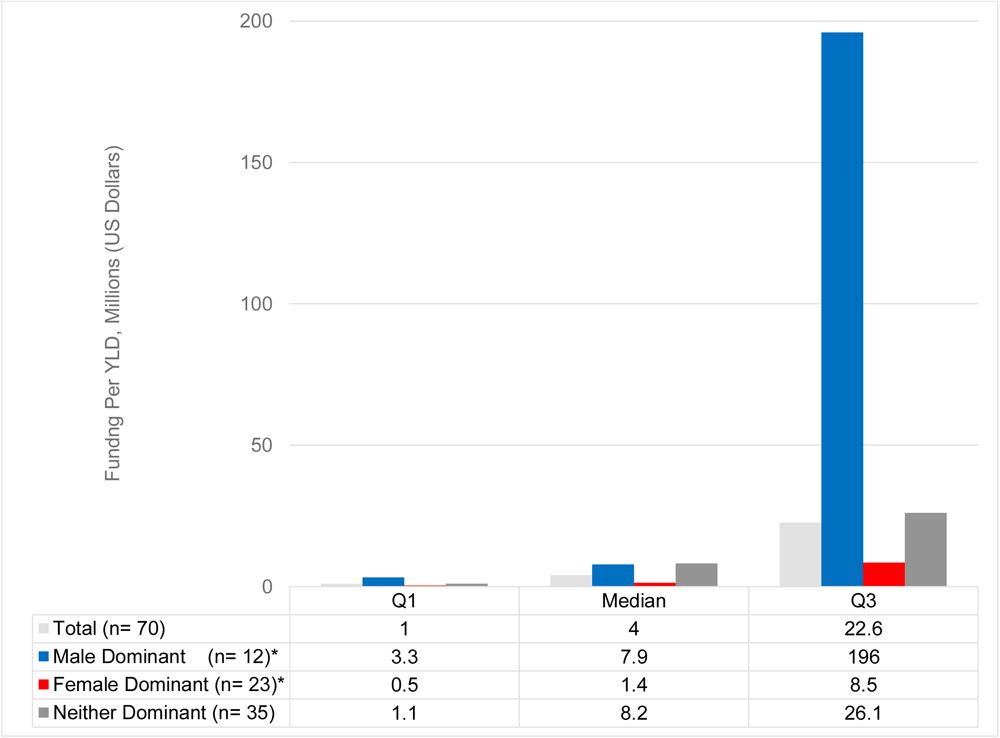
Median (IQR) Funding per YLD: Male-Female-or Neither Dominant Conditions.

## Discussion

Long COVID is a novel, disabling and likely chronic condition. Yet, there are scant data on its burden of disability, to guide scientific research and public health planning. We estimated Long COVID’s non-fatal disease burden in US adults and its actual: burden-commensurate NIH funding in 2024 relative to other conditions overall, and biological sex, Of the 70 conditions analyzed, disabling Long COVID ranked in the highest 25%ile of YLDs at 320/100,000, just above Alzheimer’s (279.4) and below asthma (355.7). Furthermore, it was among the lowest actual: burden-commensurate funded conditions at just 10% of its YLD commensurate burden. Similarly, ME/CFS, another female-dominant condition associated with up to 60% increased risk post-COVID^1^ received <1% of its actual: burden-commensurate funding.

Long COVID is “…a very insidious, beneath-the-radar-screen public health emergency…. [F]unction is being considerably impaired…[but] that doesn’t attract as much attention as a death rate,” stated Dr. Anthony Fauci.^29^ Dr. Fauci’s observation from October 2022 was perspicacious. Our findings underscore the extent to which NIH investment in Long COVID diagnostic and treatment research hasn’t tracked with its disability burden. The NIH’s median funding per condition in 2024 was $203 million, surpassing Long COVID’s $80 million and Long COVID’s common correlates of ME/CFS ($13 million) and POTS ($4 million). Admittedly, our analysis excludes the $1.15 billion in NIH RECOVER funds allocated since 2020. However, as pointed out by advocates and researchers, much of this went towards observational (vs. intervention) studies.^20^

Previously, Ballreich et.al. found no association between disease burden and NIH funding.^21^ Mirin, whose ME/CFS research informed ours, noted that only through happenstance could a merit-driven proposal review system yield a portfolio reflective of disease burden. He posited that conditions with less (vs. more) mature knowledge bases don’t fare as well with reviewers.^24^ Long COVID advocates and researchers have highlighted the need to fund studies that incorporate knowledge from other post-viral conditions (e.g., ME/CFS, dysautonomas) and patients’ lived experiences.^1,15,16^ In a positive step, the NIH allocated $10 million for an Office of Autoimmune Disease Research; 80% of autoimmune disease occur in women (https://orwh.od.nih.gov/OADR-ORWH#card-1244).^31^ While this may benefit Long COVID, ME/CFS and dysautonomias, which have autoimmune components, it does not match the burden of disease identified in our study.

Increased funding of Long COVID research furthers the NIH mission of reducing health disparities. NIH recently designated people with disabilities as a population with health disparities, joining groups such as racial/ethnic minorities, lower socioeconomic status individuals, rural populations, and sexual and gender minorities. (https://www.nih.gov/news-events/news-releases/nih-designates-people-disabilities-population-health-disparities) In recommending this designation, emphasis was placed on disability’s intersectionality with various conditions and identities associated with health disparities.^18^ Relevantly, RECOVER studies have found that Black and Hispanic Americans are more likely than white Americans to have Long COVID symptoms. Additionally, underserved, rural, vulnerable and minority populations were less likely to receive an early Long COVID diagnosis.^3,30^ And, women are consistently >=50% more likely to experience Long COVID than men.

Our findings about research funding disparities for female-predominant conditions confirm prior work. One reason may be, as discussed earlier, that because they generate psychological distress, they are presumed to be psychosomatic. ^1–3,13,23^ An oft-cited 2020 paper by Mirin found that actual vs. DALY-commensurate NIH funding was nearly twice as large for male-versus female-dominant conditions.^24^ We found more than a 5-fold difference favoring males, perhaps reflecting our use of YLDs as the burden metric, rather than DALYs. A 2023 paper in *Nature* depicts this disparity visually, while showing that if funding for women’s health were increased, there would be a huge impact on quality of life and productivity.^31^

Our study has limitations. First, harmonizing a disability weight for Long COVID from one source (IHME/GBD) with symptoms from another (HPS) introduces complexities (see “Long COVID Disease Burden”). YLDs are a function of frequency and severity. Thus, we conservatively estimated “frequency” by restricting our case definition to only the most significant activity limitations. Second, YLDs are often applied to chronic illnesses where prevalence is relatively stable over time. Further investigation is needed to determine whether and how Long COVD prevalence and disease burden changes with time. Third, our scatterplot and burden commensurate results might look different with comparator conditions other than the 70 we analyzed (because YLD estimates were available for them). Finally, as an experimental survey designed for repeated, rapid deployment, the HPS may be subject to coverage and non-response bias.^19^

Our study has multiple strengths. Paramount among them, it meets data needs identified by LC clinician-researchers: *“Accurately identifying PCC [sic; “LC”] and PCC-related burden is instrumental to inform health care policy and public health resource allocation*.”^3^ We present the first LC *disability* burden data, i.e., excluding COVID’s active phase or mortality. Now that disability accounts for most COVID disease burden,^25^ our findings are especially timely. Second, data from repeated cross-sectional survey of population-based samples increases the robustness of our findings. Third, LC qualifies as a disability in the US; the guidance accompanying this designation lists the exact symptoms we used to define disabling LC. This means our findings are directly applicable to US health care policy and public health planning.

## Conclusion

As estimated 3.8 million US adults are living with the debilitating effects of a condition that was unknown until a few years ago. The non-fatal disease burden of Long COVID is in the top 25%ile of 70 conditions we analyzed. To the extent that the subset of conditions in our study are representative of non-fatal disease burden across the NIH portfolio, our findings support designated funding, such as was established for HIV/AIDS with passage of the Ryan White HIV/AIDS program legislation in 1991. In the US, legislation has been introduced in the 2023-2024 session that will create patient registries, conduct research on post-viral disease treatments, conduct public education, and expand legal and other services In addition, our findings underscore the need for Long COVID to becoming fully recognized (vs. qualifying) as a disability in the US. Finally, by characterizing the scope of disability burden in the US, we hope this research promotes parallel population-based studies in other countries.

## Data Availability

All data produced in the present study are available upon reasonable request to the authors

