## Supplement Tables 1-3 for "Long COVID Disability Burden in US Adults: YLDs and NIH Funding Relative to Other Conditions"

### Supplement Table 1: Evidence Review

| Sample Setting, Size | Data/ Source(s) | Prevalence: Overall & Symptoms | Functional Measure: Disability, Activity, Quality of Life (QOL) |
|---|---|---|---|
| Centers for Disease Control and Prevention/ National Center for Health Statistics<br>Long Covid; Household Pulse Survey (website) Link | | | |
| ○ National Center for Health Statistics (NCHS) (items) + US Census (sampling frame) | ○ 6/2022<br>○ Online - text & email recruit<br>○ Panels<br>○ Experimental | Long Covid Definition: Covid >= 3 months prior;<br>*Specific queries range of Long Covid symptoms, e.g., fatigue, brain fog. As of April 2023:*<br>● Ever Had Long Covid:<br>○ % All US adults: 15.1%<br>○ % who ever had Covid: 27.8%<br>● Currently Have Long Covid:<br>○ % All US adults: 5.6%<br>○ % who ever had Covid: 10.1% | Activity Limitations from Long Covid, as of April 2023<br>● Among All US Adults:<br>○ Any activity limits 27.8%<br>○ Significant limits 4.6 %<br>● Among Adults Who Currently Have Long Covid:<br>○ Any activity limits: 79%%<br>○ Significant limits: 23.6%= |
| Ford et. al., 2023[1]<br>Long COVID and Significant Activity Limitation Among Adults, by Age — United States, June 1–13, 2022, to June 7–19, 2023 | | | |
| ○ National Center for Health Statistics (NCHS) (items) + US Census (sampling frame)<br>○ Subset w Current | ○ 6/2022-6/2023<br>○ Online - text & email recruit<br>○ Panels<br>○ Experimental | Long Covid Definition: Covid >= 3 months prior, June 2023<br>● Among Adults w Current LC:<br>○ Limited "A lot " 26.4% (95% CI: 24.0-8.9) | Persistence of Significant Activity Limitations from Long Covid,<br>Prevalence- of significant activity lits- stable over 1 year study period (-0.05% change per cycle, p=0.72) |
| Lau et. al., 2023.[2]<br>Physical and Mental Health Disability Associated with Long-Covid: Baseline Results From a US Nationwide Cohort. LINK | | | |
| ● Hopkins Study (US)<br>● 18+ years<br>● Hospital + Non-Hospital<br>● Baseline Status:<br>○ Long Covid: 7,926<br>○ Resolved: 948<br>○ Never: 633 | ● 1/2020-6/2022<br>● Prospective<br>● Online survey<br>● Social media & community | Long Covid Definition: Covid >= 3 months prior | Long Covid Group:<br>● 65% had >= 1 disability<br>● Mobility (difficult: walk ¼ mile or 10 steps) = 41%<br>● IADL- some difficulty= 57%<br>● Mental fatigue, severe= 6.7% |
| Ma et. al., 2023:[3]<br>Long-Term Consequences of Asymptomatic SARS-CoV-2 Infection: A Systematic Review and Meta-Analysis. Link | | | |
| ● 3 Countries- not US*<br>● 941* (adults)<br>● Criteria: >= 3 month follow up<br>● *Asymptomatic*- no symptoms at time of (+) test<br>● *Symptomatic*- not defined | ● 2020 & 2021†<br>● Observational Cohort Studies | Long Covid Definition: unstated, but criteria for study entry: >= 3-month follow-up<br>Prevalence- *Asymptomatic* Adults:<br>● >= 1 Symptom. 21%<br>● Fatigue. 15%<br>● Muscle/Joint. 9%<br>● Dyspnea. 9%<br>Asymptomatic v Symptomatic:<br>● OR for >= 1 Symptom: 0.80 | NA |
| Ma et. al., 2022: [4]<br>Long-Term Consequences of Covid 19 at 6 Months and Above: A Systematic Review and Meta-Analysis. Link | | | |
| ● Global<br>● 10,945 cases/40 studies<br>● Criteria: >=6-month Follow-up | ● < 2/9/2022<br>● Observational cohort studies | Long Covid Definition: >=6-month post-PCR confirmed Covid*<br>Prevalence: 6-12 months/ > 12 months:<br>● Long Covid: 64% / 59%<br>● Fatigue/muscle weak: 54% / 34% | Impact on Quality of Life:<br>@ 6-12 months/ @> 12 months:<br>● Pain/discomfort 33% / 35%<br>● Usual activity 7%/11%<br>● Personal care 1% /1% |

| | | | |
|---|---|---|---|
| • NA: age, hospital, symptomatic | | • Dyspnea: 75% / 81%<br>• Anxious/depressed: 33% / 35% | • Mobility 10% / 9% |
| Perlis et. al. ,2022:[5]<br>Prevalence and Correlates of Long Covid Symptoms Among US Adults. Link | | | |
| • Covid States Project- 8 waves (US)<br>• 16,091; had Covid | • 2/2021-7/2022<br>• Non-probability sample<br>• Internet sample | Long Covid Definition: Covid >= 2 months prior<br>Prevalence= 14.7%<br>US Est. Prevalence= 13.9%<br>See Table: fatigue (52%), brain fog or memory (46%s), shortness of breath (40%) | NA |
| **Sample Setting, Size** | **Data/ Source(s)** | **Prevalence: Overall & Symptoms** | **Functional Measure: Disability, Activity, Quality of Life (QOL)** |
| Rahmati et. al. 2023:[6]<br>Systematic review & meta-analysis of long-term sequelae of COVID-19 2-year after infection. Link | | | |
| • Global/12 studies<br>• Total n= 1, 289,044<br>• Criteria:<br>○ >= 2 years post-covid<br>○ >=1 outcome: heart GI, skin, respiratory, neurologic, psych<br>• Adults 18+<br>• Hospital rate: 2%-100% | • Cohort studies | Long Covid Definition: NA; >= 2 years post<br>Prevalence: 41.7%. (>=1 symptom)<br>Symptom @ 2 years: 2-year v 1-year:<br>Fatigue- 27%. Same*<br>Sleep- 25%. Better<br>Dyspnea- 10%. Same<br>Anxiety- 9%. No data<br>Focus- 8% No data<br>Myalgia- 6%. Same | NA |
| Robertson et. al. 2023:[7]<br>The Epidemiology of Long Coronavirus Disease in US Adults Link | | | |
| • US<br>• N=3,024<br>• English & Spanish | • June/July 2022<br>• Survey<br>• 64% landline<br>• Census-weighted<br>• Age & Gender- adjusted | Long Covid Definition: Covid >= 4 weeks prior + fatigue, concentration and/or shortness of breath<br>Prevalence= 7.3%* (n= 222)<br>US Estimate Prevalence= 18.8 mil<br><br>*Pulse 7/22 data= 7.5%* | Limits on (ADLs) among n=222 with Long Covid (est. US prevalence):<br>• A lot: 25% (4.8 mil)<br>• A little: 50% (9.3 mil)<br>• None: 25% (4.7 mil)<br><br>*Pulse ADL data, Sept-Dec 22: 79% *any limit* & 25% *significant* limit among those with current Long Covid* |
| Smith M., 2022:[8]<br>Estimating Total Morbidity Burden Of COVID-19: Relative Importance Of Death And Disability | | | |
| • NA: Burden models based on estimates<br>• No data collected | NA | ○ Long Covid Definition: NA, but cites Covid & ME/CFS: fatigue, muscle/body ache & difficulty concentrating. | ○ Model 1 treats Long Covid like ME/CFS & thus applies ME/CFS Disability weights per prior work:<br>○ Mild CFS: 0.14<br>○ Moderate CFS: 0.45<br>○ Severe CFS: 0.76 |
| Woodrow et.al. 2023[9]<br>Systematic Review of the Prevalence of Long Covid. Link | | | |
| • China, Europe, NA, Others<br>• 120 studies: Cohort, Cross-Sectional, Case-Control<br>• Community + Hospital + Both | • 1/2020-11/2022<br>• Systematic Review<br>• Adults & Children | Long Covid Definition: IF >= 3 months self-report of COVID or (+) test:<br>• Persistent symptoms, and/or<br>• Functional disability, and/or<br>• New pathology<br>Prevalence: Overall: 42% (95% CI: 7%-88%)<br>• Fatigue: 22%<br>• Breathing problems:15% | Function:<br>• Not Full Health/Fitness: 35% (8%-70%)<br><br>Overall- wide ranges, somewhat higher among self-reported |
| Wulf-Hanson & Vos. 2022:[10] | | | |

| Estimated Global Proportions of Individuals with Persistent Fatigue, Cognitive, and Respiratory Symptom Clusters Following <u>Symptomatic</u> COVID-19 in 2020 and 2021. [Link] | | | |
|---|---|---|---|
| ○ 22 countries<br>○ 54 studies<br>○ 1.2 million medical records<br>○ Symptomatic only | ○ 3/2020-1/2022<br>○ Medical Records<br>○ Hospitalized & Non-Hospitalized<br>○ <20 years & >=20 years | <u>Long Covid Definition</u>: Covid >= 3 months prior<br><u>Global Prevalence</u>: 6.2%<br>○ Fatigue cluster- 3.2%<br>○ Cognitive cluster- 2.2%<br>○ Respiratory cluster- 3.7%<br><u># of Symptom Clusters</u><br>○ 1 Symptom= 61.6%<br>○ 2 Symptoms= 24.4%<br>○ 3 Symptoms= 8.5%<br>Females: app.2x as MALES | <u>Disability Weight By Level:</u><br>(mild/moderate/severe)<br>• Fatigue: 0.22 (1 level)<br>• Cognitive: 0.07/0.38 (mild/severe)<br>• Respiratory: 0.02/0.23/0.41<br><br>Authors note that on average, living with Long Covid is associated with losing 21% of health.[11] |


1. Ford ND, Slaughter D, Edwards D, et al. Long COVID and Significant Activity Limitation Among Adults, by Age - United States, June 1-13, 2022, to June 7-19, 2023. *MMWR Morb Mortal Wkly Rep.* 2023;72(32):866-870.
2. Lau B, Wentz E, Ni Z, et al. Physical Health and Mental Fatigue Disability Associated with Long COVID: Baseline Results from a US Nationwide Cohort. *Am J Med.* 2023.
3. Ma Y, Deng J, Liu Q, Du M, Liu M, Liu J. Long-Term Consequences of Asymptomatic SARS-CoV-2 Infection: A Systematic Review and Meta-Analysis. *Int J Environ Res Public Health.* 2023;20(2).
4. Ma Y, Deng J, Liu Q, Du M, Liu M, Liu J. Long-Term Consequences of COVID-19 at 6 Months and Above: A Systematic Review and Meta-Analysis. *Int J Environ Res Public Health.* 2022;19(11).
5. Perlis RH, Santillana M, Ognyanova K, et al. Prevalence and Correlates of Long COVID Symptoms Among US Adults. *JAMA Netw Open.* 2022;5(10):e2238804.
6. Rahmati M, Udeh R, Yon DK, et al. A systematic review and meta-analysis of long-term sequelae of COVID-19 2-year after SARS-CoV-2 infection: A call to action for neurological, physical, and psychological sciences. *J Med Virol.* 2023;95(6):e28852.
7. Robertson MM, Qasmieh SA, Kulkarni SG, et al. The Epidemiology of Long Coronavirus Disease in US Adults. *Clin Infect Dis.* 2023;76(9):1636-1645.
8. Smith MP. Estimating total morbidity burden of COVID-19: relative importance of death and disability. *J Clin Epidemiol.* 2022;142:54-59.
9. Woodrow M, Carey C, Ziauddeen N, et al. Systematic Review of the Prevalence of Long COVID. *Open Forum Infect Dis.* 2023;10(7):ofad233.
10. Wulf Hanson S, Abbafati C, Aerts JG, et al. Estimated Global Proportions of Individuals With Persistent Fatigue, Cognitive, and Respiratory Symptom Clusters Following Symptomatic COVID-19 in 2020 and 2021. *JAMA.* 2022;328(16):1604-1615.
11. National Academies of Engineering SaM. Long COVID Examining Long-Term Health Effects of COVID-19 and Implications for the Social Security Administration: Proceedings of a Workshop. 2022.

**Supplement Table 2: LC Definitions**

Comparison of Long Covid Symptoms: Global Burden of Disease vs. Household Pulse Survey

**Global Burden of Disease**- Health states, symptom descriptions and disability weight for the 3 Long Covid symptom clusters identified by the GBD Long Covid Study*

**Table 1. Health States, Symptom Descriptions, and Disability Weights Used for the 3 Long COVID Symptom Clusters**

| Symptom cluster | Health state | Symptom description | Disability weight (95% UI)[a] |
|---|---|---|---|
| Ongoing respiratory problems | | | |
| Mild symptoms | Mild chronic respiratory problems | Cough and shortness of breath after heavy physical activity, but able to walk long distances and climb stairs | 0.02 (0.01-0.04) |
| Moderate symptoms | Moderate chronic respiratory problems | Cough, wheezing, and shortness of breath even after light physical activity; feel tired and can only walk short distances or climb a few stairs | 0.23 (0.15-0.31) |
| Severe symptoms | Severe chronic respiratory problems | Cough, wheezing, and shortness of breath all the time; great difficulty walking even short distances or climbing any stairs, feel tired when at rest, and have anxiety | 0.41 (0.27-0.56) |
| Cognitive problems | | | |
| Mild symptoms[b] | Mild cognitive problems | Some trouble remembering recent events and find it hard to concentrate and make decisions and plans | 0.07 (0.05-0.10) |
| Severe symptoms[b] | Moderate cognitive problems | Memory problems and confusion, feel disoriented, hear voices sometimes that are not real, and need help with some daily activities | 0.38 (0.25-0.51) |
| Persistent fatigue with bodily pain or mood swings | Postacute consequences of an infectious disease | Always tired and easily upset; feel pain all over the body and have depression | 0.22 (0.15-0.31) |

Abbreviation: UI, uncertainty interval.

[a] Quantifies health loss as a fraction of time lived within a health state (0 indicates full health; 1 indicates complete loss of health).

[b] Also used in the Global Burden of Disease Study for mild and moderate dementia. Additional details appear in eSection 1 in Supplement 1.

*Reprinted from: Wulf Hanson S, Abbafati C, Aerts JG, Al-Aly Z, Ashbaugh C, Ballouz T, et al. Estimated Global Proportions of Individuals With Persistent Fatigue, Cognitive, and Respiratory Symptom Clusters Following Symptomatic COVID-19 in 2020 and 2021. *JAMA*. 2022;328(16):1604-15.

**Household Pulse Survey**- identifies US adults with Long Covid by asking whether they had any of the following symptoms for 3 months or longer that they hadn't had prior to COVID-19: "

Yes" (vs. "No") The HPS identified these symptoms as including the following:

- Tiredness or fatigue,
- difficulty thinking, concentrating, forgetfulness, or memory problems (sometimes referred to as "brain fog"),
- difficulty breathing or shortness of breath,
- joint or muscle pain,
- fast-beating or pounding heart (also known as heart palpitations),
- chest pain,
- dizziness on standing,
- menstrual changes,
- changes to taste/smell, or
- inability to exercise

These symptoms are listed in the US Dept. of Health and Human Services in its Guidance on "Long Covid" as a Disability Under the ADA, Section 504, and Section 1557."

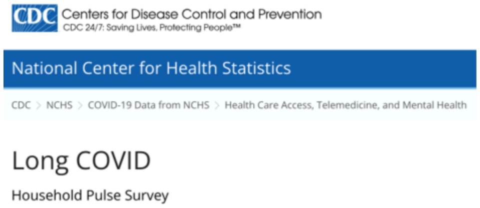

### Supplement Table 3: YLD and Funding for 70 Conditions

| NIH Research, Condition, Disease Categorization (RCDC) | YLD/ 100,000 | NIH 2024 $, millions | NIH Research, Condition, Disease Categorization (RCDC) | YLD/ 100,000 | NIH 2024 $, millions |
|---|---|---|---|---|---|
| Alcoholism, Alcohol Use and Health | 303.7 | 639 | Infertility | 8.4 | 199 |
| Alzheimer's Disease | 279.4 | 3515 | Inflammatory Bowel Disease | 43.6 | 206 |
| Anorexia | 23.4 | 16 | Kidney Disease | 232.2 | 708 |
| Anxiety Disorders | 675.7 | 274 | Liver Cancer | 2.8 | 978 |
| Arthritis | 888.1 | 324 | **Long COVID** | 319.9 | 80 |
| Asthma | 355.7 | 306 | Lung | 48.1 | 2491 |
| Attention Deficit Disorder (ADD) | 11.3 | 87 | Lung cancer | 25.1 | 555 |
| Autism | 80.9 | 341 | Lymphoma | 18.5 | 347 |
| Brain Cancer | 3.7 | 456 | Macular Degeneration | 6.6 | 104 |
| Breast Cancer | 77.5 | 850 | Malaria | 0.0 | 225 |
| Cervical Cancer | 3.7 | 171 | ME/CFS | 598.0 | 13 |
| Chronic Liver Disease and Cirrhosis | 5.6 | 978 | Methamphetamine | 18.6 | 110 |
| Chronic Obstructive Pulmonary Disease | 775.3 | 150 | Migraines | 739.6 | 49 |
| Colo-Rectal Cancer | 48.7 | 401 | Multiple Sclerosis | 40.3 | 129 |
| Dental/Oral and Craniofacial Disease | 467.8 | 754 | Nutrition | 38.2 | 2341 |
| Depression | 927.2 | 702 | Otitis Media | 11.8 | 11 |
| Diabetes | 1,188.7 | 1292 | Ovarian Cancer | 5.5 | 204 |
| Digestive Diseases | 159.3 | 2708 | Pancreatic Cancer | 4.6 | 307 |
| Digestive Diseases - (Gallbladder) | 80.2 | 22 | Parkinson's Disease | 33.8 | 277 |
| Digestive Diseases - (Peptic Ulcer) | 9.0 | 9 | Perinatal Period, Conditions Originating in | 158.0 | 940 |
| Down Syndrome | 2.0 | 143 | Pneumonia | 5.6 | 160 |
| Drug Abuse (NIDA only) | 1,140.5 | 1663 | Prescription Drug Abuse | 964.5 | 129 |
| Eating Disorders | 85.6 | 61 | Preterm, Low Birth Weight, Newborn Health | 123.2 | 458 |
| Endometriosis | 21.1 | 29 | Prostate Cancer | 86.9 | 322 |
| Epilepsy | 74.7 | 244 | Psoriasis | 127.6 | 15 |
| Fibroid Tumors (Uterine) | 25.4 | 16 | Rheumatoid Arthritis | 81.3 | 95 |
| Headaches | 833.3 | 58 | Schizophrenia | 397.5 | 311 |
| Heart Disease - Coronary Heart Disease | 120.4 | 424 | Sexually Transmitted Infections | 17.3 | 395 |
| Hepatitis | 2.6 | 378 | Sickle Cell Disease | 5.5 | 161 |
| Hepatitis - A | 2.1 | 6 | Stroke | 433.5 | 444 |
| Hepatitis - B | 0.2 | 138 | Substance Abuse | 1,444.2 | 2597 |
| Hepatitis - C | 0.1 | 113 | Suicide | 18.4 | 174 |
| HIV/AIDS | 54.2 | 3294 | Tuberculosis | 1.5 | 635 |
| Hodgkin's Disease | 2.4 | 21 | Urologic Diseases | 49.0 | 682 |
| Hypertension | 47.9 | 402 | Uterine Cancer | 16.6 | 42 |
| | | | Violence Research | 81.1 | 194 |
